# Genome-wide association studies of *LRRK2* modifiers of Parkinson’s disease

**DOI:** 10.1101/2020.12.14.20224378

**Authors:** Dongbing Lai, Babak Alipanahi, Pierre Fontanillas, Tae-Hwi Schwantes-An, Jan Aasly, Roy N. Alcalay, Gary W. Beecham, Daniela Berg, Susan Bressman, Alexis Brice, Kathrin Brockman, Lorraine Clark, Mark Cookson, Sayantan Das, Vivianna Van Deerlin, Matthew Farrer, Joanne Trinh, Thomas Gasser, Stefano Goldwurm, Emil Gustavsson, Christine Klein, Anthony E. Lang, J. William Langston, Jeanne Latourelle, Timothy Lynch, Karen Marder, Connie Marras, Eden R. Martin, Cory Y. McLean, Helen Mejia-Santana, Eric Molho, Richard H. Myers, Karen Nuytemans, Laurie Ozelius, Haydeh Payami, Deborah Raymond, Ekaterina Rogaeva, Michael P. Rogers, Owen A. Ross, Ali Samii, Rachel Saunders-Pullman, Birgitt Schüle, Claudia Schulte, William K. Scott, Caroline Tanner, Eduardo Tolosa, James E. Tomkins, Dolores Vilas, John Q. Trojanowski, The 23andMe Research Team, Ryan Uitti, Jeffery M. Vance, Naomi P. Visanji, Zbigniew K. Wszolek, Cyrus P. Zabetian, Anat Mirelman, Nir Giladi, Avi Orr Urtreger, Paul Cannon, Brian Fiske, Tatiana Foroud

**Affiliations:** Department of Medical and Molecular Genetics, Indiana University School of Medicine, Indianapolis, IN, U.S.A.; 23andMe, Inc., Sunnyvale, CA, U.S.A.; Google LLC., Palo Alto, CA, U.S.A.; Department of Neurology, St. Olavs hospital, Trondheim, Norway; Department of Neurology, Columbia University, New York, NY, U.S.A.; John P. Hussman Institute for Human Genomics and Dr. John T. Macdonald Department of Human Genetics, University of Miami, Miller School of Medicine, Miami, FL, U.S.A.; Department of Neurology, Christian-Albrechts-University of Kiel, Germany; Department of Neurodegenerative Diseases, Hertie Institute for Clinical Brain Research, University of Tübingen, Tübingen, Germany; Department of Neurology, Icahn School of Medicine at Mount Sinai, New York, NY, U.S.A.; Sorbonne Université, Institut du Cerveau et de la Moelle épinière (ICM), AP-HP, Inserm, CNRS, University Hospital Pitié-Salpêtrière, Paris, France; German Center for Neurodegenerative Diseases (DZNE), Tübingen, Germany; Department of Pathology and Cell Biology, Columbia University, New York, NY, U.S.A.; Laboratory of Neurogenetics, National Institute of Aging, National Institute of Health, Bethesda, MD, U.S.A.; Department of Pathology and Laboratory Medicine, University of Pennsylvania, Philadelphia, PA, U.S.A.; Fixel Institute for Neurological Diseases, McKnight Brain Institute, L5-101D, UF Clinical and Translational Science Institute, University of Florida, FL, U.S.A.; Institute of Neurogenetics, University of Luebeck, Luebeck, Germany; Parkinson Institute, ASST “G.Pini-CTO”, Milan, Italy; Centre for Applied Neurogenetics, University of British Columbia, Vancouver, British Columbia, Canada; The Edmond J Safra Program in Parkinson Disease and the Morton and Gloria Shulman Movement Disorders Clinic, Toronto Western Hospital, 399, Bathurst St, Toronto, ON, M5T 2S8, Canada; Departments of Neurology, Neuroscience, and of Pathology, Stanford University School of Medicine, Stanford, CA, U.S.A.; GNS Healthcare, Cambridge, MA, USA; Dublin Neurological Institute at the Mater Misericordiae University Hospital, Conway Institute of Biomolecular and Biomedical Research, University College Dublin, Dublin, Ireland; Department of Neurology, Psychiatry, Taub Institute and Sergievsky Center, Columbia University Vagelos College of Physicians And Surgeons, New York, NY, U.S.A.; The Edmond J Safra Program in Parkinson’s disease, Toronto Western Hospital, University of Toronto, Toronto, Canada; Google LLC., Cambridge, MA, U.S.A.; Gertrude H. Sergievsky Center, Columbia University, New York, NY, U.S.A.; Department of Neurology, Albany Medical College, Albany, NY, U.S.A.; Department of Neurology, Boston University, Boston, MA, U.S.A.; Department of Neurology, University of Alabama at Birmingham, Birmingham, AL, U.S.A.; Tanz Centre for Research in Neurodegenerative Diseases and Department of Neurology, University of Toronto, Toronto, Ontario, Canada; Department of General Surgery, University of South Florida Morsani College of Medicine, Tampa, FL, U.S.A.; Departments of Neuroscience and Clinical Genomics, Mayo Clinic, Jacksonville, FL, U.S.A.; School of Medicine and Medical Science, University College Dublin, Dublin, Ireland; VA Puget Sound Health Care System and Department of Neurology, University of Washington, Seattle, WA, U.S.A.; Department of Pathology, Stanford University School of Medicine, Stanford, CA, U.S.A.; University of California, San Francisco Veterans Affairs Health Care System, San Francisco, CA, U.S.A.; Parkinson disease and Movement Disorders Unit, Hospital Clínic Universitari, Institut d’Investigacions Biomèdiques August Pi i Sunyer (IDIBAPS), University of Barcelona (UB), Centro de Investigación Biomédica en Red sobre Enfermedades Neurodegenerativas (CIBERNED), Barcelona,Spain; School of Pharmacy, University of Reading, Reading, Berkshire, UK; Department of Neurology, Mayo Clinic, Jacksonville, Florida; Tel Aviv Sourasky Medical Center, Sackler Faculty of Medicine and Sagol School of Neuroscience, Tel Aviv University, Tel Aviv, Israel; The Michael J. Fox Foundation for Parkinson’s Research, NY, U.S.A.

**Author notes:** Corresponding Author Dongbing Lai, Ph.D., 410 W. 10^th^ Street, HS 4000, HITS, Indianapolis, IN 46202-3002. These authors have equal contributions (Dongbing Lai, Babak Alipanahi, Pierre Fontanillas).

## Abstract

**Objective:** The aim of this study was to search for genes/variants that modify the effect of *LRRK2* mutations in terms of penetrance and age-at-onset of Parkinson’s disease.

**Methods:** We performed the first genome-wide association study of penetrance and age-at-onset of Parkinson’s disease in *LRRK2* mutation carriers (776 cases and 1,103 non-cases at their last evaluation). Cox proportional hazard models and linear mixed models were used to identify modifiers of penetrance and age-at-onset of *LRRK2* mutations, respectively. We also investigated whether a polygenic risk score derived from a published genome-wide association study of Parkinson’s disease was able to explain variability in penetrance and age-at-onset in *LRRK2* mutation carriers.

**Results:** A variant located in the intronic region of *CORO1C* on chromosome 12 (rs77395454; P-value=2.5E-08, beta=1.27, SE=0.23, risk allele: C) met genome-wide significance for the penetrance model. A region on chromosome 3, within a previously reported linkage peak for Parkinson’s disease susceptibility, showed suggestive associations in both models (penetrance top variant: P-value=1.1E-07; age-at-onset top variant: P-value=9.3E-07). A polygenic risk score derived from publicly available Parkinson’s disease summary statistics was a significant predictor of penetrance, but not of age-at-onset.

**Interpretation:** This study suggests that variants within or near *CORO1C* may modify the penetrance of *LRRK2* mutations. In addition, common Parkinson’s disease associated variants collectively increase the penetrance of *LRRK2* mutations.

## Introduction

Parkinson’s disease (PD) is the second most common neurodegenerative disease in older adults.^1^ Several genes showing autosomal dominant (*SNCA, LRRK2, VPS35*) or recessive (*PRKN, PINK1, DJ-1*) inheritance patterns have been identified as the cause of familial PD. These genes harbor rare, high penetrance mutations that explain up to 10% of familial PD cases in different populations.^1,2^ Recently, large genome-wide association studies (GWAS) have identified over 90 loci with small individual effects on disease risk in both familial and sporadic PD.^3,4^

Mutations in *LRRK2* are among the most common genetic causes of PD.^1,2^ The most frequent mutation is G2019S (rs34637584), which explains up to 10% of familial PD cases and 1-2% of all PD cases.^2,5^ Among PD patients, the frequency of the G2019S mutation is approximately 3% in Europeans, 16-19% in Ashkenazi Jews and up to 42% in Arab-Berbers.^6-14^ Estimates of the risk for developing PD among *LRRK2* G2019S mutation carriers range from 15% to 85%.^15-18^ To explain the incomplete penetrance of G2019S, it has long been hypothesized that there are other genes/variants outside of *LRRK2* acting to modify its effect (*LRRK2* modifiers). Identification of *LRRK2* modifiers could aid the development of novel prevention and treatment strategies for PD.

Most studies of *LRRK2* modifiers, to date, have focused on candidate genes. Since the protein product of *LRRK2* may interact with α-synuclein (encoded by *SNCA*), and tau (encoded by *MAPT*),^19,20^ variants in *SNCA* and *MAPT* were widely investigated. However, the results have been inconsistent, possibly due to small sample sizes and differences in variants and populations investigated.^21-29^ Other PD associated genes such as *GBA*,^28^ *BST1*,^28^ *GAK*,^29^ and PARK16^28,30,31^ have also been investigated. However, the number of studies is limited and findings remain to be replicated. Genome-wide searches for *LRRK2* modifiers are sparse and limited to linkage studies. Using 85 *LRRK2* carriers from 38 families, a genome-wide linkage study of *LRRK2* modifiers found a suggestive linkage region at 1q32 (LOD=2.43); but that study did not identify any candidate genes/variants underlying the linkage peak.^32^ A genome-wide linkage scan in Arab-Berber PD families found *DNM3* as a *LRRK2* modifier.^33^ This finding was not independently replicated, although it was still significant in a meta-analysis including the participants reported in the original finding.^24,34^ Genome-wide association studies have successfully detected many disease genes/variants, including those associated with PD. However, to date, no GWAS for *LRRK2* modifiers has been reported, probably due to limitations in sample size and corresponding statistical power.

In this study, we recruited *LRRK2* mutation carriers from multiple centers and performed the first GWAS to identify genes/variants that modify the penetrance and age-at-onset of PD among *LRRK2* mutation carriers. Using the largest cohort to date, which consisted of 1,879 *LRRK2* mutation carriers (including 776 PD cases), one genome-wide significant association signal was found in the intronic region of the *CORO1C* gene. In addition, we found that a polygenic risk score (PRS) derived from publicly available PD GWAS summary statistics, was associated with penetrance, but not age-at-onset, of PD in *LRRK2* mutation carriers.

## Methods

### Study participants

The studies and the *LRRK2* mutation carriers were grouped into three cohorts. The first cohort was primarily identified from The Michael J. Fox Foundation’s *LRRK2* Consortium and consisted of research sites worldwide (referred to as the MJFF consortium cohort). We searched PubMed and identified study groups that reported *LRRK2* mutation carriers then asked them to participate in this study (PUBMED IDs: 16240353, 16333314, 18986508, and 16960813).^35-38^ We also made announcements at international conferences to recruit more study mutation carriers. Details can be found in their publications.^35-38^ To maximize participation and facilitate uniform data preparation across sites, a minimal dataset was submitted for all subjects that included *LRRK2* mutation status, sex, age-at-onset (for PD cases), age at last evaluation (for non-PD participants) and pedigree information, along with the availability of a minimal amount of DNA (∼2 ug). The minimal phenotypic data were sent to Indiana University and the subjects were assigned a unique identifier. The second cohort was from Tel Aviv University, Israel (referred to as the Israel cohort). Participants were of Ashkenazi origin and recruited from the Movement Disorders Unit at Tel Aviv Medical Center. PD diagnosis was confirmed by a movement disorders specialist and clinical disease status (PD or not diagnosed as PD) was evaluated at the time of blood draw for genetic testing. The third cohort (referred to as the 23andMe cohort) consisted of research participants of the personal genetics company 23andMe, Inc. who were *LRRK2* G2019S carriers and whose PD status was known. Individuals who reported via an online survey that they had been diagnosed with PD by a medical professional, were asked to provide their age at diagnosis. For individuals who affirmed at least once that they had not been diagnosed with PD, their age at the most recent completion of the survey was recorded. The study was approved by the Institutional review board at Indiana University; the Institutional Review Board (Helsinki) Committee of Tel Aviv Sourasky Medical Center and the National Helsinki Committee for Genetic Research in Humans, MINISTRY OF HEALTH, Israel; Ethical & Independent Review Services, a private institutional review board (http://www.eandireview.com).

### Genotyping, quality review, and imputation

All study participants were genotyped on the Illumina Omni 2.5 Exome Array V1.1 (Illumina, San Diego, USA), except 166 participants from the Israel cohort, who were genotyped on an earlier version of the same array (V1.0). This array has common, rare, and exonic variants that were selected from diverse world population samples included in the 1000 Genomes Project. In total, there were >2.58M variants, including >567K exonic variants. Participants from the MJFF consortium and 23andMe were genotyped at the Center for Inherited Disease Research (CIDR) at Johns Hopkins University (Baltimore, MD, USA). The Israel cohort was genotyped at Tel Aviv University and two samples from the MJFF consortium were included for quality control. There were 134 duplicated and unexpected identical participants among all three cohorts. Pairwise concordance rates were all >99.97%, showing high consistency among the two genotyping labs and the two versions of the Illumina array.

Variants with genotypic missing rates >5% and non-polymorphic variants were excluded. In addition, variants with A/T or C/G alleles were also excluded due to strand ambiguity. Hardy-Weinberg equilibrium (HWE) was not used to filter variants because these participants were ascertained to be *LRRK2* mutation carriers and this participant selection scheme would directly violate HWE and remove potential *LRRK2* modifiers from the analysis.

To confirm the reported pedigree structure and detect cryptic relatedness, we used a set of 56,184 high quality (missing rate <2%, HWE P-values >0.001), common (MAF >0.1), and independent (linkage disequilibrium as measured by r^2^ <0.5) variants to calculate the pairwise identity by descent using PLINK.^39^ Reported pedigree structures were revised accordingly, if necessary. Mendelian error checking was performed in the revised pedigree structure. Any inconsistent genotypes were set to missing. The same set of variants was also used to estimate the principal components (PCs) of population stratification using Eigenstrat.^40^ All samples were imputed to the Haplotype Reference Consortium (http://www.haplotype-reference-consortium.org/) using Minimac3.^41^ A total of 725,802 high quality genotyped variants were selected for imputation (MAF >3%, HWE P-value >0.0001, and missing rate <5%). EAGLE v2.4^42^ was used to phase genotyped variants for each sample. After filtering out variants with poor imputation quality score (R^2^ < 0.6) and checking for Mendelian inconsistencies using PLINK,^39^ a final dataset of 7,934,276 imputed and genotyped variants was used for association analyses.

### Genome-wide association studies

Our association analysis tested two models: 1) variants modifying the penetrance for PD among *LRRK2* mutation carriers (penetrance model), and 2) variants modifying the age-at-onset for PD among *LRRK2* mutation carriers (age-at-onset model). For the penetrance model (including PD cases and those not diagnosed as PD at last evaluation), the association analysis was designed to identify variants associated with the time to PD diagnosis or last evaluation for undiagnosed mutation carriers. For the age-at-onset model (PD cases only), the association analysis tested whether variants contributed to the age-at-onset for PD cases among *LRRK2* mutation carriers.

For the penetrance model, a mixed effect Cox proportional hazard model (frailty model) was used with sex, 10 PCs, array and cohort indicators as covariates. Family relationships were adjusted by using a kinship matrix calculated using R package COXME (https://cran.r-project.org/web/packages/coxme/index.html). For the age-at-onset model, a linear mixed model was fit with the same covariates as the penetrance model and a kinship matrix to adjust family relationships. Variants with MAF > 1% were tested for association in these two models. In addition to the single variant analyses, we performed gene-based association analyses for both the penetrance and age-at-onset models. We focused on rare exonic and splicing variants, based on annotations from Variant Effect Predictor (https://useast.ensembl.org/info/docs/tools/vep/index.html), and restricting to variants with MAF <3%. Only genotyped variants (N= 725,802) were used in the gene-based analyses due to the low quality of imputation for rare variants. The R package COXME was used to perform all analyses (https://cran.r-project.org/web/packages/coxme/index.html). Conditional analysis was conducted using the most significant variant in an associated region as a covariate, and additional signals within the associated region were determined based on P-values < 0.01.

### Functional studies

To evaluate whether rs77395454 has immediate biological consequences on gene expression (eQTL) of nearby genes, we searched Open Targets Genetics (https://genetics.opentargets.org/) and GTEx (https://www.gtexportal.org/). In addition, protein-protein interaction (PPI) data was assessed to identify whether the protein product which maps to this locus either interacts directly with LRRK2 or has common interactors that are shared with LRRK2, using PINOT (Protein Interaction Network Online Tool) version 1.0^43^ queried on 16th June 2020 (http://www.reading.ac.uk/bioinf/PINOT/PINOT_form.html).

### Polygenic risk score analyses

In the largest GWAS analysis of PD susceptibility to date, Nalls et al meta-analyzed 17 datasets with 56,306 PD cases or proxy-cases and 1.4 million controls.^4^ Based on their results, they developed a PRS using summary statistics of 1,805 variants that can explain 26% of PD heritability.^4^ In this study, we performed PRS analysis using these 1,805 variants. Detailed information about how to select these 1,805 variants was described in Nalls et al.^4^ Since we were searching for *LRRK2* modifiers, variants in the *LRRK2* region (chr12: 40,118,913-41,263,086) were excluded. The PRS was calculated as a weighted summation of effective alleles with the logarithm of odds ratios as the weights. This derived PRS was used to fit the same models with the same set of covariates as described for the genome-wide association analyses using the R package COXME.

## Results

Study participants from the three cohorts are summarized in **Table 1**. In total, 1,879 participants (853 individuals from 294 families and 1,026 singletons) were included in the analyses. Among them, 776 had, or self-reported, a PD diagnosis and 1,103 were not classified as affected with PD at the last evaluation. The majority of participants were G2019S carriers, only 4% carried other *LRRK2* mutations as reported by the contributing sites, all from the MJFF consortium cohort. In the 23andMe cohort, 85% of participants were not diagnosed with PD and most of them were less than 50 years of age at the time of their last evaluation. Based on PCs, the majority of participants were of European ancestry.

**Table 1:**
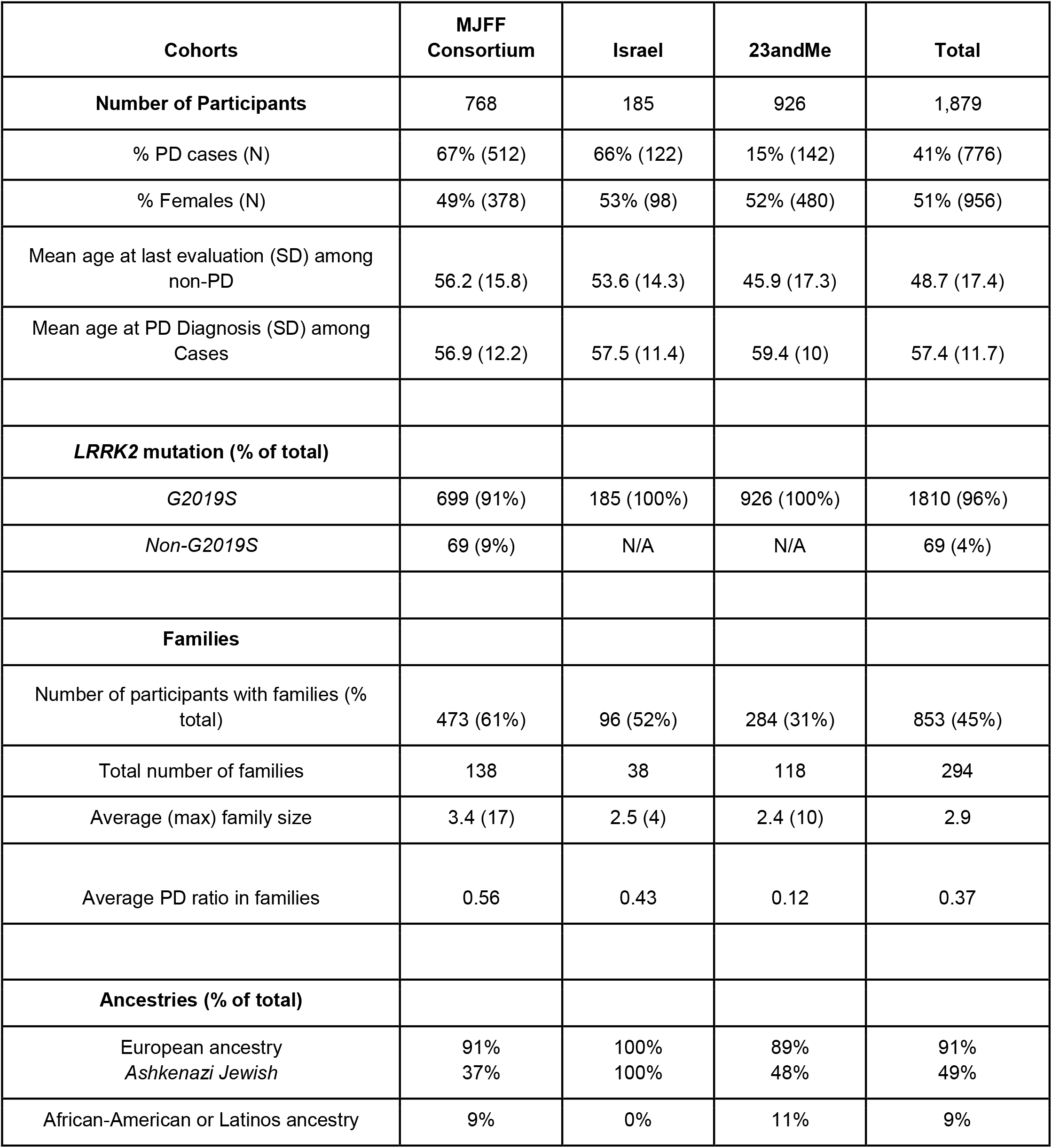
Summary of study cohorts.

Manhattan plots for the single variant analyses of the penetrance and age-at-onset models are shown in **Figure 1**. Q-Q plots for both models are show in **Figure 1C**. No obvious bias was detected in either model, genomic controls were 1.055 and 1.052 for the penetrance model and age-at-onset model, respectively. Twelve loci showed variants with P-values <1.0E-6 (i.e. meet the threshold for suggestive significance) in either the penetrance model or the age-at-onset model (**Table 2)**. One variant on chromosome 12 reached genome-wide significance (rs77395454, P-value=2.5E-08) in the penetrance model. Conditional analysis suggested that there were no additional association signals in this locus. The top variant (rs77395454) on the chromosome 12 region is located in an intron of *CORO1C* (coronin 1C) (**Figure 2**). The causal haplotype(s) spanned *SELPLG, CORO1C*, and *SSH1*, with most of the variants within *CORO1C*. **Figures 3A, 3B, and 3C** show the survival curves stratified by rs77395454 genotypes for all samples, familial samples, and unrelated samples, respectively. Heterozygous rs77395454 carriers (20 familial and 37 unrelated samples) had an increased risk of PD. Six other loci met suggestive significance (P-value< 1.0E-6) for the penetrance model (**Table 2**).

**Table 2:** Variants that have P-values < 1.0E-6 in either penetrance model or age-at-onset model.

|  |  |  |  |  |  |  | Penetrance model |  |  |  | Age-at-onset model |  |  |  |
| --- | --- | --- | --- | --- | --- | --- | --- | --- | --- | --- | --- | --- | --- | --- |
| CHR | BP | rsid | Alleles | Gene | Annotation | MAF | BETA | SE | P-Value | G2019S only P-value | BETA | SE | P-Value | G2019S only P-value |
| 1 | 221,173,137 | rs141686162 | A/G | *HLX,DUSP10* | Intergenic | 0.01 | 0.42 | 0.28 | 0.13 | 0.86 | 1.33 | 0.27 | 8.5E-07 | 1.1E-04 |
| 3 | 124,083,400 | rs145611031 | C/G | *KALRN* | Intron | 0.02 | 1.14 | 0.23 | 5.2E-07 | 2.3E-07 | 0.98 | 0.23 | 2.3E-05 | 9.1E-06 |
| 3 | 140,288,373 | rs150382576 | A/G | *CLSTN2* | 3'UTR | 0.02 | 0.78 | 0.22 | 5.3E-04 | 2.1E-03 | 1.19 | 0.23 | 2.7E-07 | 8.7E-07 |
| 3 | 152,841,926 | rs59679443 | A/G | *RAP2B,A RHGEF26* | Intergenic | 0.04 | 0.62 | 0.17 | 3.1E-04 | 1.6E-04 | 0.86 | 0.18 | 9.3E-07 | 1.1E-06 |
| 3 | 152,932,435 | rs16846845 | G/C | *RAP2B,A RHGEF26* | Intergenic | 0.05 | 0.83 | 0.16 | 1.1E-07 | **4.5E-08** | 0.72 | 0.16 | 9.4E-06 | 1.0E-06 |
| 4 | 160,854,320 | rs12272007 | A/G | *LOC107986324* | Intergenic | 0.01 | 1.22 | 0.36 | 6.0E-04 | 1.9E-03 | 1.81 | 0.36 | 3.6E-07 | 3.6E-06 |
| 5 | 115,786,384 | rs73781088 | C/T | *SEMA6A* | Intron | 0.03 | 0.47 | 0.19 | 0.01 | 0.05 | 1.00 | 0.20 | 6.8E-07 | 5.1E-07 |
| 8 | 9,520,115 | rs28398294 | G/A | *TNKS* | Intron | 0.03 | 1.09 | 0.22 | 5.7E-07 | 1.1E-06 | 0.76 | 0.22 | 4.8E-04 | 1.1E-03 |
| 9 | 127,532,973 | rs148922482 | C/T | *NR6A1* | Intron | 0.01 | 0.90 | 0.28 | 1.4E-03 | 3.7E-03 | 1.40 | 0.28 | 6.3E-07 | 2.5E-06 |
| 11 | 120,585,515 | rs28470321 | G/A | *GRIK4* | Intron | 0.01 | 1.91 | 0.36 | 9.0E-08 | 6.1E-08 | 1.77 | 0.34 | 2.1E-07 | 1.4E-07 |
| 12 | **109,080,567** | **rs77395454** | **C/T** | **CORO1C** | **Intron** | **0.02** | **1.27** | **0.23** | **2.5E-08** | **1.0E-06** | **0.68** | **0.25** | **6.2E-03** | **0.48** |
| 14 | 90,982,388 | rs76788674 | A/G | *CALM1,TT C7B* | Intergenic | 0.03 | 0.78 | 0.16 | 7.1E-07 | 2.8E-06 | 0.57 | 0.17 | 7.6E-04 | 2.6E-03 |
| X | 123,652,525 | rs185981774 | A/G | *TENM1* | Intron | 0.02 | 0.86 | 0.17 | 5.4E-07 | 3.9E-07 | 0.62 | 0.18 | 6.0E-04 | 5.6E-04 |

Note: Genome-wide significant variant is in bold.

**Figure 1:**
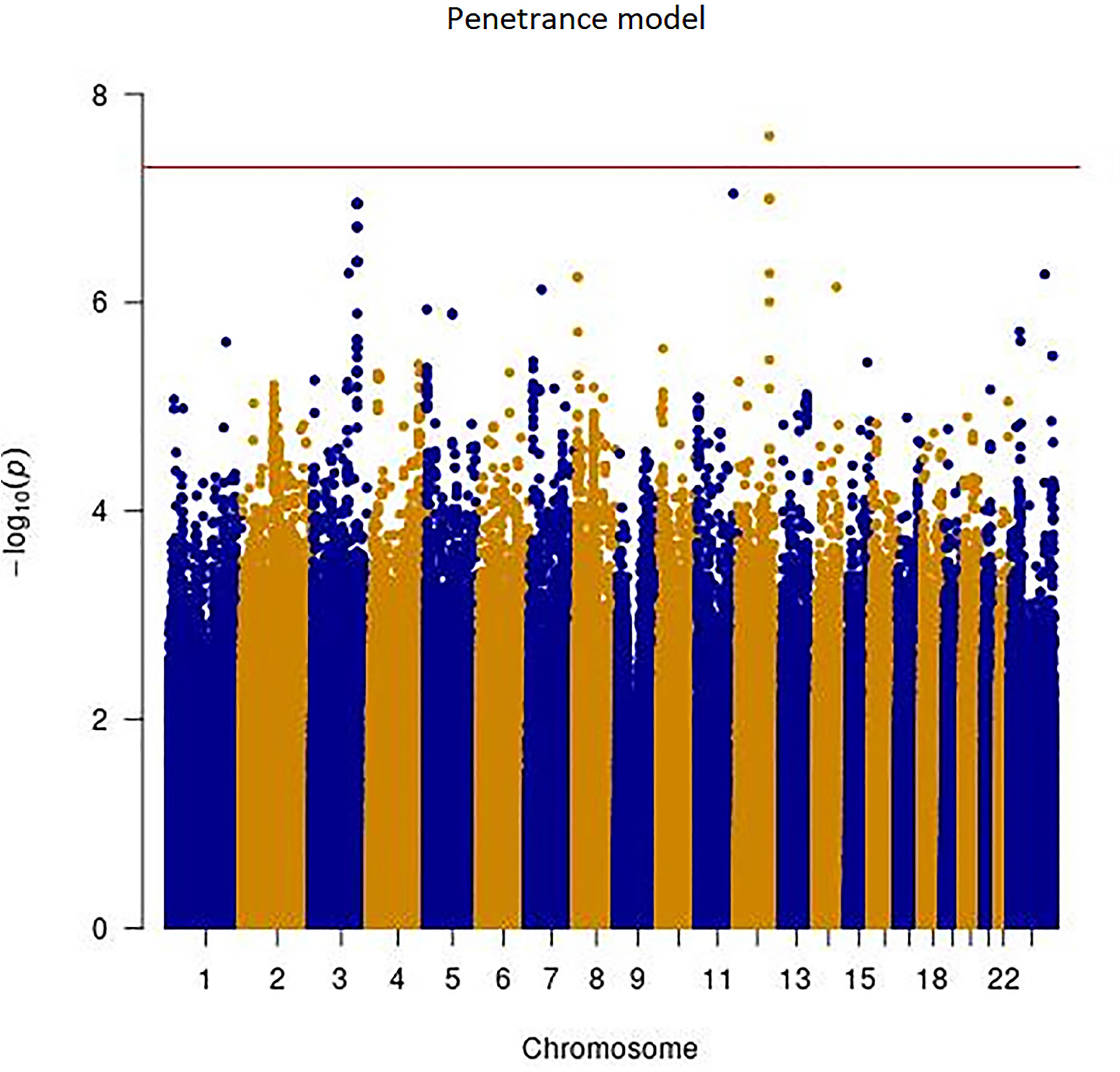
Manhattan and Q-Q plots of single variant analysis of penetrance and age-at-onset models. Y-axis is the –log(p-value) for associations. X-axis is physical position of the variants across the genome. The horizontal line indicates genome-wide significance. A: penetrance model; B: age-at-onset model; C: Q-Q plots of penetrance model (left) and age-at-onset model (right)

**Figure 2:**
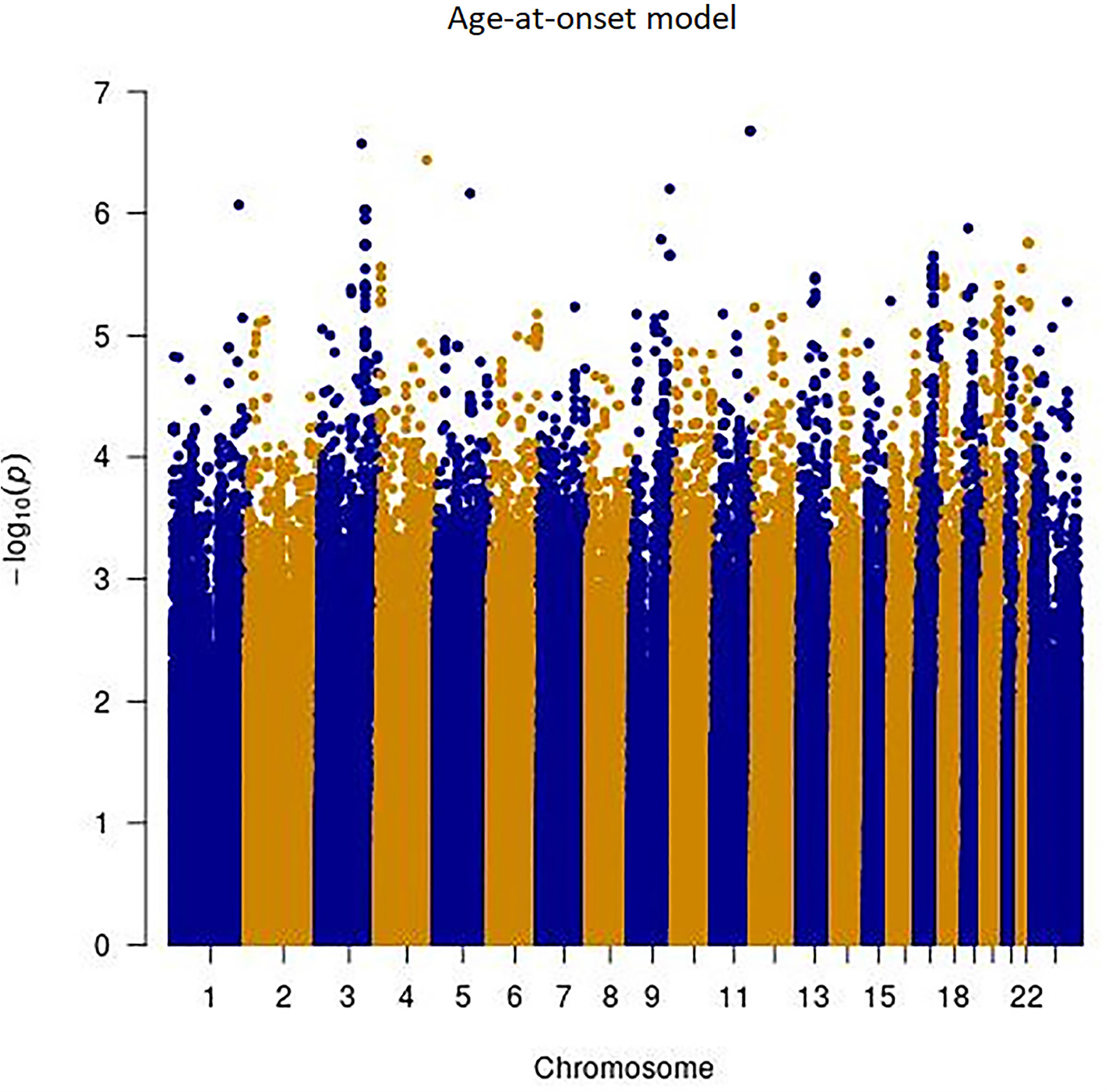
Regional association plot of the chromosome 12 region for the penetrance model. Y-axis is the -log(p-value) for associations. X-axis denotes physical positions on the chromosome (Mb). The color scale shows the extent of linkage disequilibrium (LD, as measured by r^2^) between each variant and the top variant (indicated by the purple diamond) with larger r^2^ indicating greater LD. Peaks indicate the recombination hot spots.

**Figure 3:**
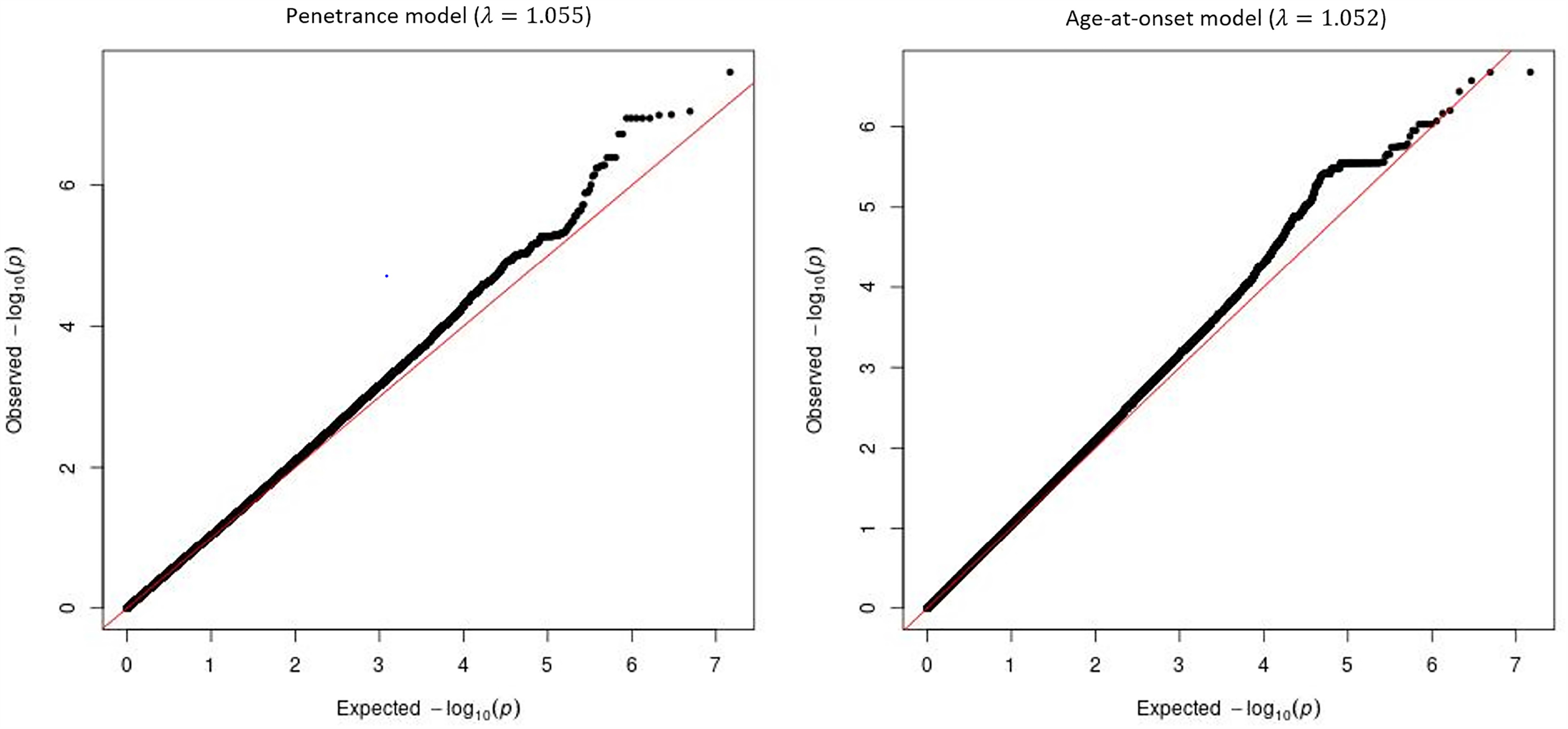
Cumulative incidence of PD stratified by rs77395454 genotypes. Dashed lines indicate 95% confidence interval. Due to the low MAF of rs77395454 and therefore the small number of CC genotype carriers, only participants with TT and CT genotypes are shown.

For the age-at-onset model, no chromosomal region reached genome-wide significance, but seven loci met the suggestive association threshold. Except for variants on chromosome 3 identified in both models, rs73781088 on chromosome 5 (intron of *SEMA6A*) for the age-at-onset model and rs28398284 on chromosome 8 (intron of *TNKS*) for the penetrance model, all other variants had no or marginal LD support. Variants on chromosome 3 from both models cover the same region but identified different haplotypes. For comparison purposes, we also performed analyses limited to only the *LRRK2* G2019S carriers; the results were comparable (**Table 1**). In addition, we performed analyses using only individuals of predicted Ashkenazi Jewish ancestry. Results are less significant due to dramatically decreased sample sizes as shown in **Supplemental Table 1**. No genome-wide significant results were detected in the gene-based analysis using exonic variants for either model.

By searching Open Targets Genetics and GTEx, we found that the most significant variant, rs77395454, is an eQTL of *CORO1C* in blood and *MYO1H* in visceral adipose (omentum). The minor allele (C allele) is associated with higher expression of *CORO1C* and *MYO1H*. We did not find evidence that LRRK2 interacts with CORO1C or MYO1H directly. However, there are several proteins that are the common interactors of both LRRK2 and CORO1C: ABCE1, ACTR2, CDC42, DAPK1, MYO1C, RAC1, and TP53, identified using PINOT.^43^

Among those 1,805 variants that were obtained from the study of Nalls et al 2019,^4^ 20 variants were not present in our datasets. An additional 27 variants were located in the *LRRK2* region and were excluded, and 1,758 variants were included in the PRS calculation. The PRS was a significant predictor in the penetrance model (P-value=7.8E-4) but not in the age-at-onset model (P-value=0.75). These results suggest that a high genetic risk of PD significantly increases the chance of developing PD among *LRRK2* mutation carriers.

## Discussion

Two major unresolved questions in PD research are why some, but not all, *LRRK2* mutation carriers develop PD, and why the age-at-onset is so variable in those that do. This work represents the first GWAS study to report *LRRK2* modifiers of PD penetrance and age-at-onset. One variant on chromosome 12 reached genome-wide significance in the penetrance model (rs77395454 in an intronic region of *CORO1C*). Several loci reached suggestive significance in either the penetrance model or the age-at-onset model. One region on chromosome 3 showed suggestive associations in both models. PRS derived from a publicly available PD GWAS was a significant predictor of penetrance of PD among *LRRK2* mutation carriers.

The genome-wide significant variant, rs77395454 on chromosome 12, is located in an intronic region of *CORO1C*. There are several proteins that are common interactors of both LRRK2 and CORO1C. Two of them, CDC42 and RAC1, have previously been validated as modifiers of *LRRK2*-mediated neurite shortening (reviewed in Boon et al., 2014).^44^ These results suggest that both CORO1C and LRRK2 might have effects on the actin cytoskeleton. Furthermore, a recent APEX2 screen identified that CORO1C is physically proximate to LRRK2 in cells.^45^ Finally, the protein expression of CORO1C is significantly higher in *LRRK2* knockout mice *in vivo*, as shown by proteomics and validated by western blotting.^46^ The accumulation of CORO1C in knockout mice might represent compensation for diminished *LRRK2* function. The CORO1C protein is a member of the WD repeat protein family, which has been implicated in signal transduction and gene regulation (https://www.ncbi.nlm.nih.gov/gene/23603). In a zebrafish model of spinal muscular atrophy, over-expression of *CORO1C* rescued the phenotype caused by *SMN* deficiency.^47^ Using mass spectrometry, Malty et al. showed that the product of *CORO1C* interacts with mitochondrial proteins associated with neurodegeneration.^48^ Collectively, these complementary results suggest that *CORO1C* is a more likely functional interactor of *LRRK2*. However, it is possible that other genes in this region may underpin the observed association. For example, the protein product of *SSH1* regulates actin filament dynamics, which has been linked to *LRRK2* mutations.^49,50^ *SELPLG* has been linked to neuropsychiatric disorders such as conduct disorder.^51^ Further studies are needed to conclusively determine the gene(s) underlying the observed association.

Multiple variants on chromosome 3 were supported by both models, although they identified different associated haplotypes. The most significant variants were rs16846845 in the penetrance model and rs150382576 in the age-at-onset model. This region is under a known linkage peak for PD (LOD=2.5).^52^ In the study by Gao et al., two variants (rs902432 and rs755763) had LOD scores >2 in different analysis models.^52^ These two variants are about 850Kb upstream and 200Kb downstream from variants identified in our study, respectively. This is consistent with our findings that top variants in either model, and variants in LD with them, were physically distinct from each other. A nearby region was also linked to PD (LOD=3.6) in an Amish Parkinsonism pedigree linkage study performed by Lee at al.^53^ In both the Gao et al and Lee et al linkage studies, no candidate genes were nominated due to the large size of the reported linkage regions.^52,53^ Variants that we identified are located near *RAP2B*, a member of the *RAS* oncogene family. However, its role in PD is unknown. Rs73781088 on chromosome 5 is in the intronic region of *SEMA6A*, which is broadly expressed in the brain. This gene is associated with amyotrophic lateral sclerosis.^54^ Rs28398294 on chromosome 8 is in the intronic region of *TNKS*, which is also broadly expressed in the brain. This region has been linked to Alzheimer’s disease.^55^ Rs141686162 on chromosome 1 is in an intergenic region near *DUSP10*, which has been associated with progressive supranuclear palsy in a recent study.^56^ All of these findings warrant further study to investigate their potential roles in modifying the effect of *LRRK2* mutations.

We also examined the variants previously reported as *LRRK2* modifiers in other studies. Thirteen variants from seven genes passed our QC (rs4273468 from *BST1*; rs2421947 from *DNM3*; rs1564282 from *GAK*; rs1052553, rs242562, and rs2435207 from *MAPT*; rs823144 from *PARK16*; rs11931074, rs1372525, rs181489, rs2583988, and rs356219 from *SNCA*; rs11578699 from *VAMP4*).^19-31,33,34^ Only four variants from three genes had P-values < 0.05: rs823144 from *PARK16* in the penetrance model (P-value=0.01); rs1564282 from *GAK* in both the penetrance (P-value=0.03) and age-at-onset models (P-value=7.1E-03); rs2345207 (P-value=5.1E-04) and rs1052553 (P-value=0.02) from *MAPT* in the penetrance model. Unfortunately, since some individuals in our study may have also been included in the previous studies where these candidate genes were first reported, our findings do not represent independent replication. However, our results showed that previously reported variants on *BST1, DNM3, SNCA*, and *VAMP4* were not replicated and they are likely not *LRRK2* modifiers.

The significant effect of the PRS in the penetrance model supports the polygenic nature of the *LRRK2* modifiers, i.e. there are many genetic variants each with a small effect that collectively have a significant effect on the risk of PD in *LRRK2* mutation carriers. This result is in line with the recent analysis of Iwaki et al.^57^ In that study, a PRS was derived using 89 genome-wide significant variants (some of which were also included in our PRS) identified in a PD GWAS of Nalls et al.^4^. Iwaki et al. found that the PRS was significantly associated with the penetrance of the *LRRK2* G2019S mutation. Potential overlap between the participants in our study and that of Iwaki et al. means that the results of these studies do not represent independent replication. We did not detect a significant association in the age-at-onset model. One reason for this may be the smaller sample size (less than half of that in the penetrance model, only 776 affected from 1,879 total participants analyzed), and the resulting lack of statistical power. Another possible reason could be that the PRS was derived from a GWAS comparing PD cases and controls, and these risk associated genes/variants are not necessarily associated with age-at-onset.

There are several limitations of this study. First, despite the effort to enroll as many participants as possible, the sample size of this study still resulted in only modest statistical power. With this sample size, assuming a linear model, for a variant with MAF 3%, a change of at least six years of age-at-onset can be detected with 80% power at a genome-wide significant level. Second, to maximize the number of eligible studies to join this collaboration, we required a minimal set of inclusion criteria. While this approach dramatically increased the sample size, many potentially important covariates were not collected; therefore, we could not adjust for all relevant covariates in our analyses. Third, approximately 96% of our participants were G2019S carriers. However, there are carriers of other *LRRK2* mutations in the MJFF cohort. Although in a sensitivity analysis using only G2019S carriers, we observed similar effects for those top variants that we identified in both models, these mutations may still have different effects that cannot be detected in the small number of carriers. Fourth, our study cohorts consisted of family participants and unrelated participants. Family history was not collected for every participant. Therefore, some unrelated individuals may be sporadic PD and have different penetrance from familial PD participants. Fifth, there was a lack of information on subjects with subtle signs of PD but who did not yet merit a diagnosis of PD. Nevertheless, we detected a genome-wide significant variant and the PRS analysis suggested that there is unlikely to be one or several single *LRRK2* modifiers, but similar to overall PD risk, penetrance of *LRRK2* mutations is affected by multiple genetic variants.

Given the significant therapeutic efforts underway to develop targets for PD patients carrying *LRRK2* mutations, further replication of these results is essential. Furthermore, the genetic variants identified in this study and the PRS evaluated in the *LRRK2* mutation carriers, may be used in the future to make personalized prevention and treatment possible.

## Data Availability

Aggregate-level data included in this study will be made available to qualified investigators upon request. Investigators interested in receiving 23andMe data, either alone or in combination with data from other cohorts, will need to sign a Data Transfer Agreement with 23andMe that protects 23andMe research participant privacy, and should visit https://research.23andme.com/dataset-access/ to submit a request.

## Acknowledgments

We thank the research participants from all sites who made this study possible. Members of the 23andMe Research Team are: Michelle Agee, Adam Auton, Robert K. Bell, Katarzyna Bryc, Sarah L. Elson, Nicholas A. Furlotte, David A. Hinds, Karen E. Huber, Aaron Kleinman, Nadia K. Litterman, Matthew H. McIntyre, Joanna L. Mountain, Elizabeth S. Noblin, Carrie A.M. Northover, Steven J. Pitts, J. Fah Sathirapongsasuti, Olga V. Sazonova, Janie F. Shelton, Suyash Shringarpure, Chao Tian, Joyce Y. Tung, Vladimir Vacic, and Catherine H. Wilson. We thank Tara Candido for collecting the data. The authors acknowledge the Indiana University Pervasive Technology Institute for providing [HPC (Big Red II, Karst, Carbonate)] visualization, database, storage, or consulting resources that have contributed to the research results reported within this paper.

This work is supported by The Michael J. Fox Foundation for Parkinson’s Research Grants 7984, 7984.01, 7984.02 and 8981. Enrollment and clinical characterization of participants was in part funded by grants from the NIH (R01 NS065070, P50 NS062684) and the Department of Veterans Affairs (5I01CX001702).

The cohort from Columbia University, New York, NY, was funded by the Parkinson’s Foundation, the NIH (K02NS080915 and UL1 TR000040), and the Brookdale Foundation. The Mayo Clinic was a Morris K. Udall Parkinson’s Disease Research Center of Excellence (NINDS P50 NS072187) and is an American Parkinson Disease Association Information and Referral Center. This study at the Mayo Clinic was also partially funded by the gifts from The Sol Goldman Charitable Trust, the Haworth Family Professorship in Neurodegenerative Diseases fund, and The Albertson Parkinson’s Research Foundation. This work was supported in part by NINDS R01-NS078086, NINDS Tau Center without Walls (U54-NS100693 and U54-NS110435), DOD award (W81XWH-17-1-0249), The Michael J. Fox Foundation, the Little Family Foundation, the Functional Genomics of LBD Program, the Mangurian Foundation Lewy Body Dementia Program at Mayo Clinic and Mayo Clinic Center for Individualized Medicine.

The University of Miami Parkinson disease research group was a Morris K. Udall Parkinson’s Disease Research Center of Excellence (NINDS P50 NS039764 and NS071674). Genotyping services were provided by the Center for Inherited Disease Research (CIDR). CIDR is fully funded through a federal contract from the National Institutes of Health to The Johns Hopkins University, contract number HHSN268201200008I.

R.N.A received research funding from the NIH, the Parkinson’s Foundation, and The Michael J. Fox Foundation. V.V.D is supported by NS053488, AG010124, and AG062418.

T.F is supported by R01NS096740 and The Michael J. Fox Foundation.

C.K. received funding from the DFG (FOR2488).

E.M. is supported by the Riley Chair in Parkinson’s Disease.

K.M. is supported by NIH NS036630, UL1TR001873, the Parkinson’s Disease Foundation, The Michael J. Fox Foundation.

E.R. was in part supported by the Canadian Consortium on Neurodegeneration in Aging.

J.Trojanowski. is supported by AG010124 and AG062418.

J. Trinh. received research funding from the Canadian Institutes of Health Research, Else Kroener Fresenius Foundation and DFG (FOR2488).

J.E.T. is supported by the 2019 Biomarkers Across Neurodegenerative Diseases Grant Program, BAND3 (Michael J Fox Foundation, Alzheimer’s Association, Alzheimer’s Research UK and Weston Brain Institute [grant number 18063]).

## Author Contributions

TF and PC contributed to the conception and design of the study. DL, BA, PF, PC and TF drafted the text and prepared the figures. DL, BA, PF, TS, JA, RNA, GWB, DB, SB, AB, KB, LC, MC, SD, VVD, MF, J.Trinh, TG, SG, EG, CK, AEL, JWL, JL, TL, KM, CM, ERM, CYM, HM, EM, RHM, KN, LO, HP, DR, ER, MPR, OAR, AS, RS, BS, CS, WKS, CT, ET, JET, DV, J.Trojanowski, RU, JMV, NPV, ZKW, CPZ, AM, NG, AOU and BF contributed to acquisition and analysis of data. All authors reviewed and approved the submission.

---

**Supplemental Table 1:** Results of AJ ancestry sample only.

|  |  |  |  |  |  |  | Penetrance model |  |  | Age-at-onset model |  |  |
| --- | --- | --- | --- | --- | --- | --- | --- | --- | --- | --- | --- | --- |
| CHR | BP | rsid | Alleles | Gene | Annotation | MAF | BETA | SE | P-value | BETA | SE | P-value |
| 1 | 221,173,137 | rs141686162 | A/G | *HLX,DUSP10* | Intergenic | 0.01 | 0.75 | 0.46 | 0.11 | 0.74 | 0.48 | 0.12 |
| 3 | 124,083,400 | rs145611031 | C/G | *KALRN* | Intron | 0.02 | 1.05 | 0.29 | 2.74E-04 | 0.96 | 0.29 | 9.57E-04 |
| 3 | 140,288,373 | rs150382576 | A/G | *CLSTN2* | 3'UTR | 0.02 | 0.50 | 0.25 | 0.04 | 1.11 | 0.28 | 5.46E-05 |
| 3 | 152,841,926 | rs59679443 | A/G | *RAP2B,ARHGEF26* | Intergenic | 0.04 | 0.37 | 0.20 | 0.07 | 0.60 | 0.23 | 0.01 |
| 3 | 152,932,435 | rs16846845 | G/C | *RAP2B,ARHGEF26* | Intergenic | 0.05 | 0.68 | 0.19 | 2.89E-04 | 0.77 | 0.20 | 1.47E-04 |
| 4 | 160,854,320 | rs12272007 | A/G | *LOC107986324* | Intergenic | 0.01 | 0.85 | 0.47 | 0.07 | 1.73 | 0.52 | 7.96E-04 |
| 5 | 115,786,384 | rs73781088 | C/T | *SEMA6A* | Intron | 0.03 | 0.70 | 0.26 | 5.89E-03 | 0.91 | 0.26 | 5.39E-04 |
| 8 | 9,520,115 | rs28398294 | G/A | *TNKS* | Intron | 0.03 | 1.16 | 0.23 | 3.34E-07 | 0.74 | 0.25 | 3.24E-03 |
| 9 | 127,532,973 | rs148922482 | C/T | *NR6A1* | Intron | 0.01 | 0.99 | 0.40 | 1.26E-02 | 1.00 | 0.43 | 0.02 |
| 11 | 120,585,515 | rs28470321 | G/A | *GRIK4* | Intron | 0.01 | NA | NA | NA | NA | NA | NA |
| 12 | 109,080,567 | rs77395454 | C/T | *CORO1C* | Intron | 0.02 | 1.48 | 0.53 | 5.51E-03 | 0.66 | 0.56 | 0.24 |
| 14 | 90,982,388 | rs76788674 | A/G | *CALM1,TTC7B* | Intergenic | 0.03 | 0.75 | 0.27 | 6.49E-03 | 0.78 | 0.29 | 6.82E-03 |
| X | 123,652,525 | rs185981774 | A/G | *TENM1* | Intron | 0.02 | 0.85 | 0.16 | 1.31E-07 | 0.67 | 0.17 | 8.84E-05 |

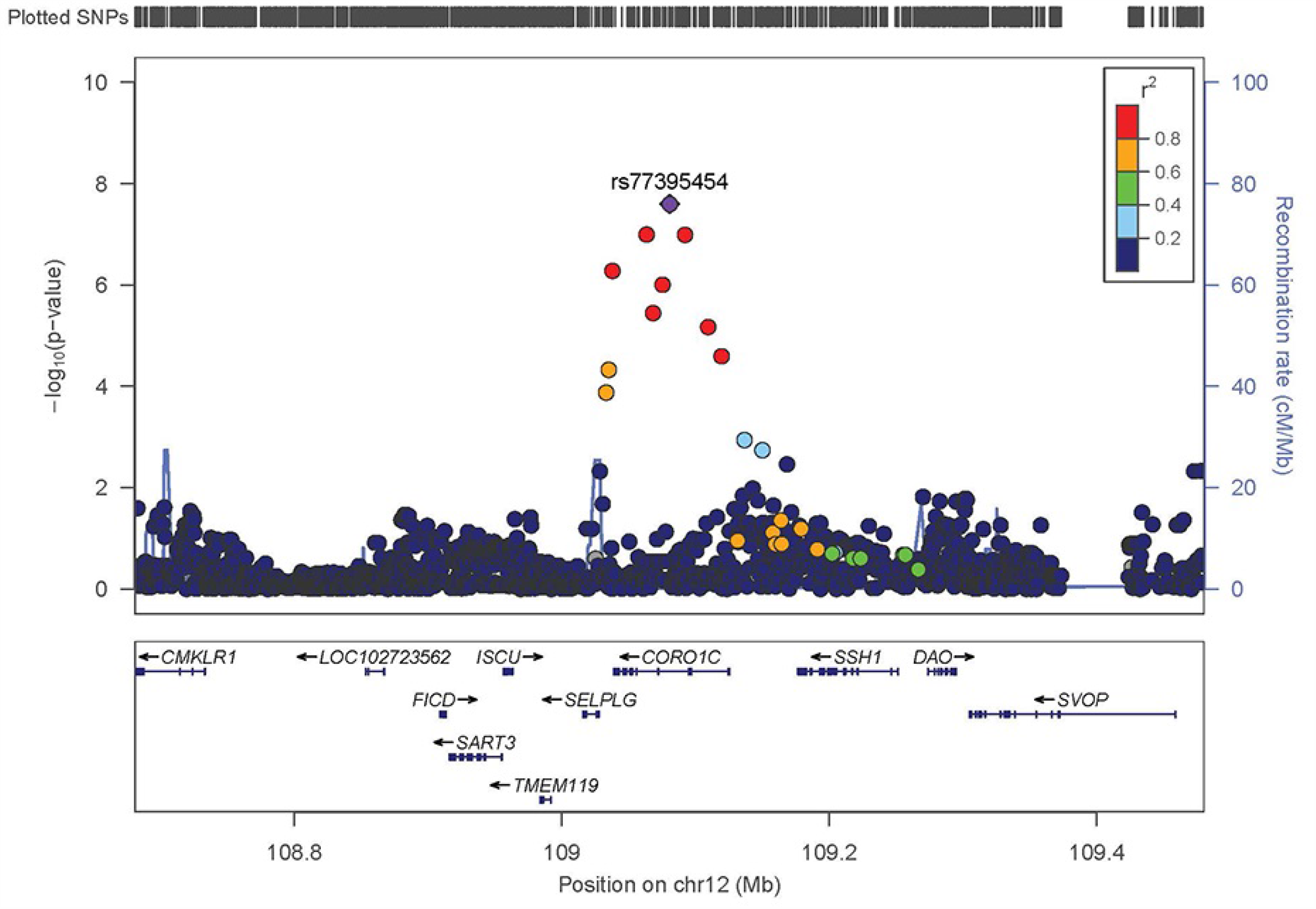

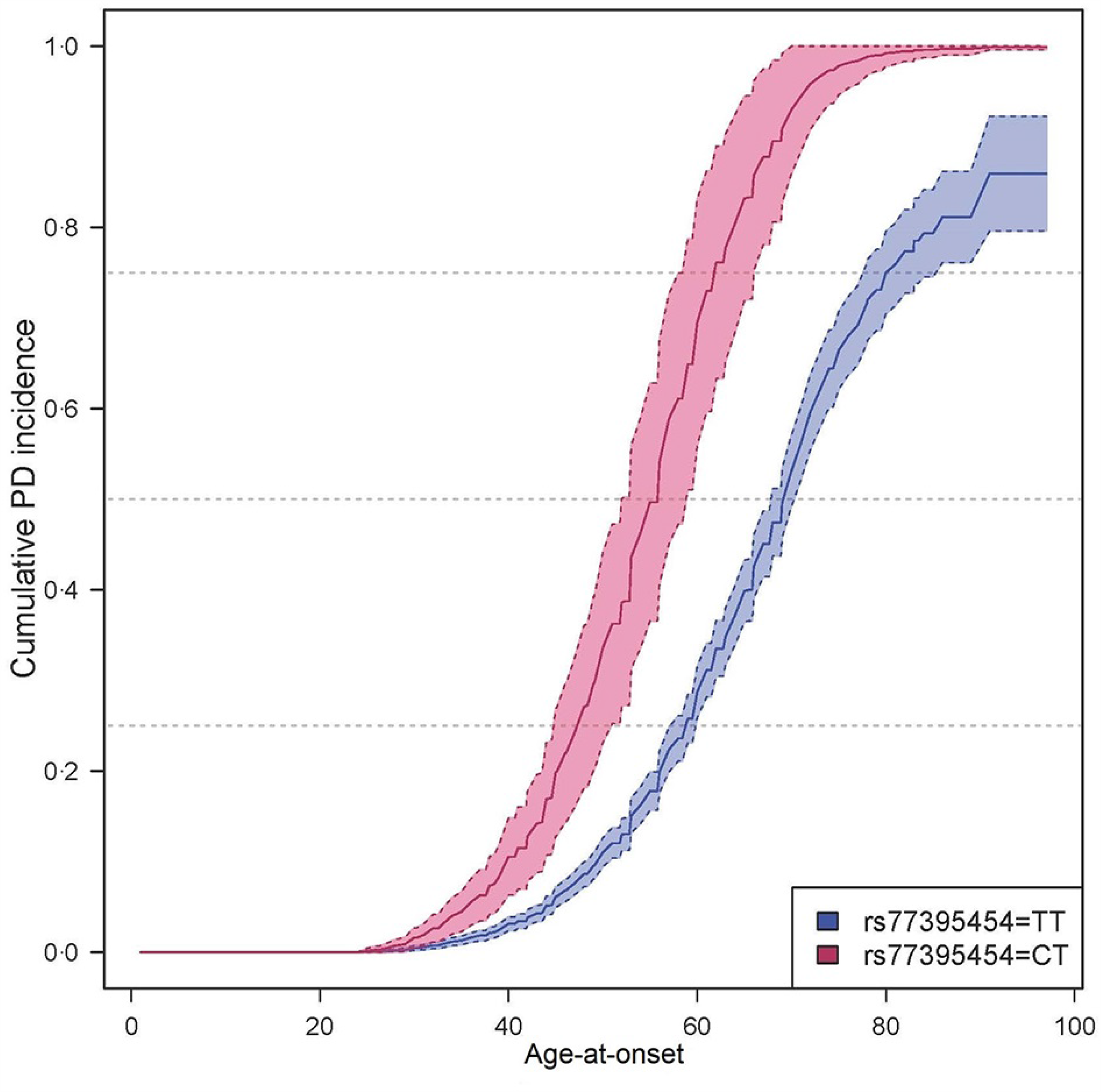

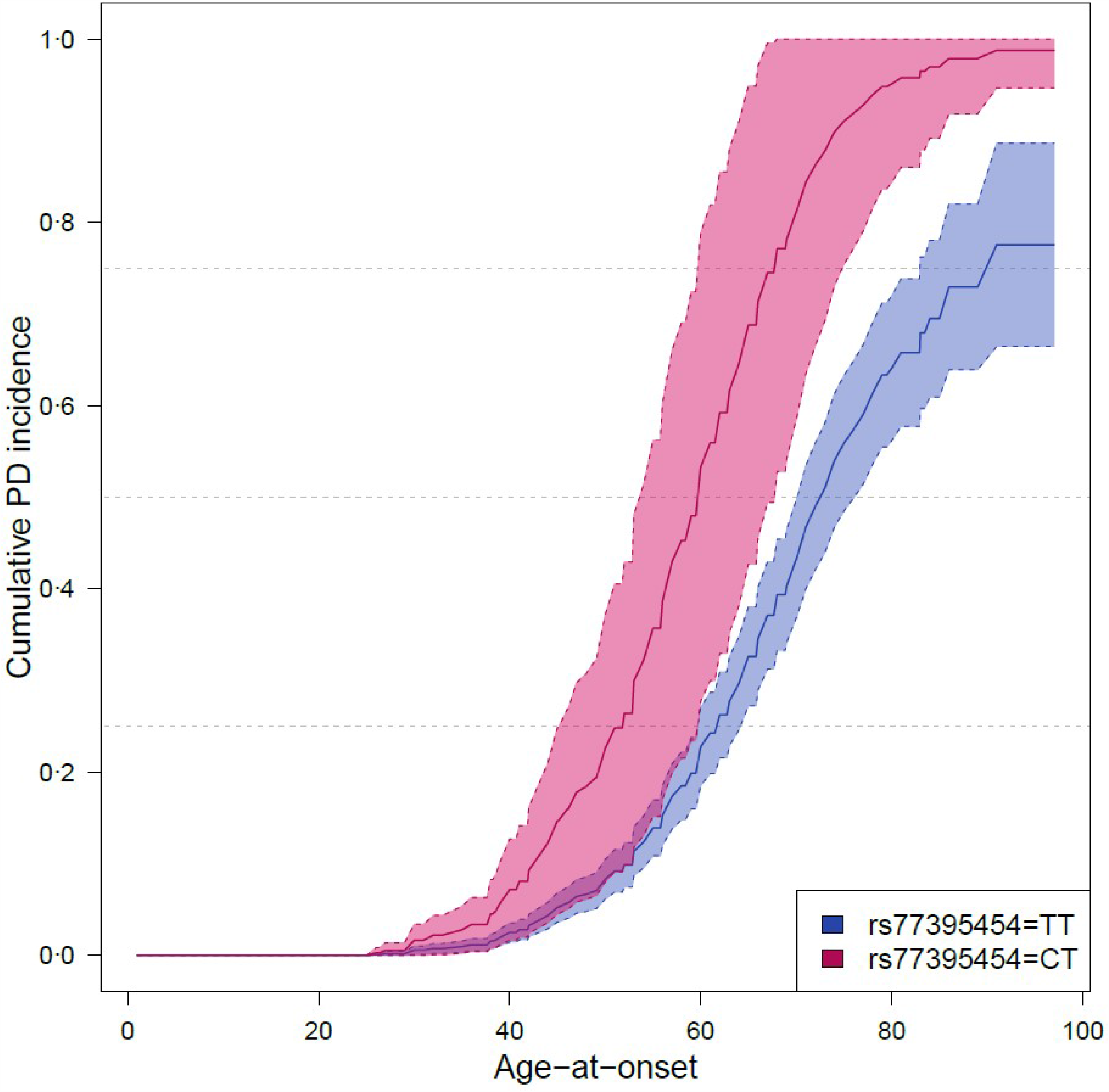

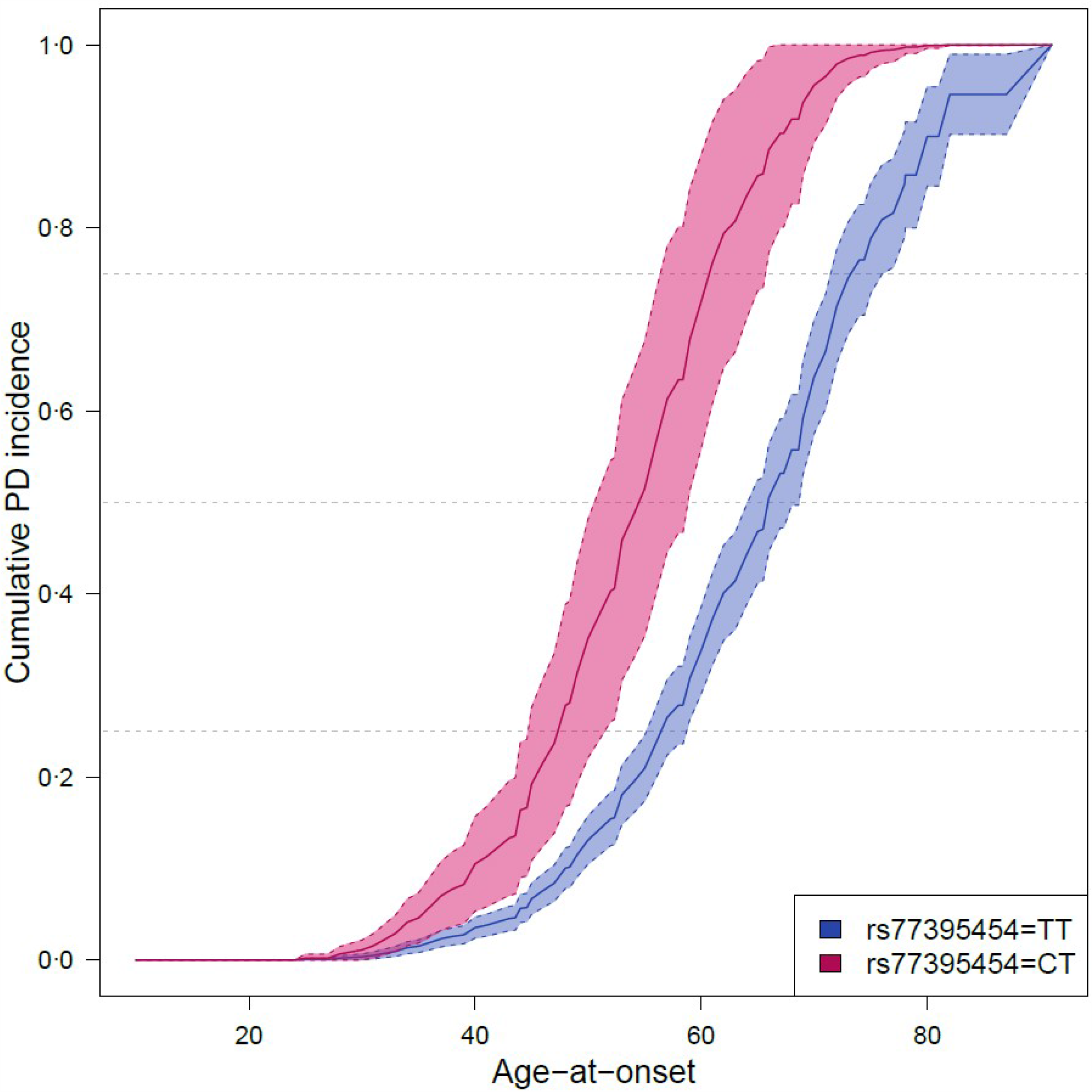

